# Willingness to Pay for Solid Waste Management Services and Associated Factors in Mbarara District, Southwestern Uganda

**DOI:** 10.1101/2025.08.26.25334428

**Authors:** Erastus Tugume, Abraham Muhwezi, Tom Murungi, Julius Kyomya, Edgar Mulogo Mugema, Richard Migisha, Moses Ntaro

**Author notes:** ***Corresponding author:*** Erastus Tugume.

## Abstract

**Background:** Solid waste management is a major environmental and public health challenge in Mbarara District, especially with rapid urbanization and limited public funding. Willingness to pay (WTP) for solid waste management services is essential for sustainability, yet it remains unassessed in the area. This study assessed the prevalence of WTP for solid waste management services and the associated factors among households in Mbarara District, Southwestern Uganda.

**Methods:** A cross-sectional quantitative study was conducted among 250 randomly selected individuals in households of Bwizibwera-Rutooma and Rubindi-Ruhumba town councils, Mbarara district. Data were collected using interviewer-administered questionnaires, entered in Microsoft Excel and transferred to STATA version 17.0 for cleaning and analysis. Continuous variables were summarized using means and standard deviations, while categorical variables were presented as frequencies and proportions. Bivariate and multivariable logistic regression analysis at a 95% level of confidence was done to identify factors associated with WTP for solid waste management services.

**Results:** Overall, 62% (156/250; 95% C.I.: 56.2%-68.2%) of the participants were willing to pay for solid waste management services. The majority, 64.1% (100/156) were willing to pay one thousand Uganda shillings or more for SWM. Factors associated with WTP for solid waste management services were; being male (aOR = 2.4, 95% CI: 1.2-4.6; *p*-value = 0.011), having a monthly income>100,000 Uganda Shillings (UGX) (aOR = 2.2, 95% CI: 1.7-4.1; *p*-value = 0.015), disposing wastes using town council services (aOR = 7.75, 95% CI: 1.35-44.47; *p*-value = 0.022) and receiving weekly waste collection services (aOR = 2.62; 95% CI: 1.06-6.50; *p*=0.038).

**Conclusion:** The willingness to pay for solid waste management services was relatively high and was positively associated with being male, monthly income>100,000 UGX, reliance on town council waste collection services and Weekly collection of the solid wastes. Solid waste management programs should ensure regular, affordable services through town councils and promote inclusive strategies that consider income and gender to enhance willingness to pay.

## Introduction

Global solid waste generation has increased significantly in recent decades due to urbanization and rising consumption (1). For example, cities worldwide produced an estimated 1.3 billion tons of municipal solid waste in 2012 (2), and solid waste generation in towns is expected to surge by 70% by the year 2050(1). This surge in waste has exceeded the capacity of many waste management systems, especially in low- and middle-income countries. In numerous developing cities, waste services already utilize 20–50% of municipal budgets, yet they often collect only a portion of the waste produced (3, 4). About 125 million tons of solid waste are generated annually across Africa, with sub-Saharan countries accounting for roughly 65% of that total, despite low collection rates (5). Uganda generates about 0.55kg/capita/day, with the collection rates being below 50% (6, 7).

The rapid urbanization and growth of cities and towns in Uganda (8) have exacerbated solid waste generation, posing significant challenges to effective solid waste management (7). This has overwhelmed the capacities of the municipal councils to manage the large quantities of waste generated (9). These municipal councils are allocated limited funds and resources to sustain waste collection services (10, 11). As a result, there has been an increase in poor solid waste management practices posing significant health risks and environmental degradation (12, 13). As a result, privatizing solid waste collection has been adopted to address poor solid waste management practices and alleviate the challenges faced by the public sector during waste collection (14-16).

Willingness to pay (WTP) for solid waste management services is a critical indicator of community support for sustainable waste systems, especially in low-resource settings. Studies across sub-Saharan Africa show considerable variation in WTP levels, with higher prevalence observed in Nigeria (64.4%)(17), Ethiopia (83.5%)(18) and Tanzania (63%)(19). However, studies in Uganda have reported WTP for solid waste management to be 48% in Lira (20) and 64% in Kawempe division, Kampala (21). The WTP levels have reportedly been influenced by factors such as income, education, household size, amount of waste generated, distance to collection sites and current service quality (17, 18, 20, 21).

With the elevation of several town councils in Mbarara District in 2019, there has been an influx of people seeking economic and social opportunities in these growing urban centres (9). This population growth, however, has brought with it significant challenges such as an increase in solid waste generation (9, 16, 22). Of the 31.423 tons of garbage produced daily across Mbarara District, the town councils of Bwizibwera-Rutooma and Rubindi-Ruhumba alone account for approximately 8 tons per day (22, 23). Yet, due to limited financial and logistical capacity, the councils are only able to collect about 60% of this waste (9, 16). Furthermore, municipal reports indicate that fewer than 30% of residents in these town councils currently pay fees for waste collection services. Given that payment for solid waste management (SWM) is increasingly recognized as a pathway toward sustainable service financing, there is a pressing need to assess the level of willingness to pay (WTP) for SWM services and the factors influencing it in these newly urbanized settings of Mbarara District. Therefore, we assessed the willingness to pay for solid waste management and associated factors among households in Mbarara district, Southwestern Uganda.

## Materials and methods

### Study design and setting

This was a cross-sectional study conducted in households of Rubindi-Ruhumba and Bwizibwera-Rutooma town councils within Mbarara district, Southwestern Uganda. The study was conducted from 11^th^ March 2025 to 14^th^ April 2025. The district is located 270km away from Kampala, the capital city of Uganda. In Mbarara district, more than 30% of the waste produced daily (over 200 tons) is left uncollected, improperly processed, or disposed of in unsuitable locations (22, 23). This has raised sanitation and health concerns in the area as poor waste disposal-related diseases have increased in the population (24, 25).

### Study population

The study was conducted among adults (≥18 years) residing in households in Rubindi-Ruhumba and Bwizibwera-Rutooma town councils in Mbarara District, who had been in the area for at least six months. Individuals with cognitive impairments or serious illnesses that hindered their ability to participate in the study were excluded.

### Sampling procedure and sample size

Out of 5 Town councils in Mbarara district, 2 Town councils were randomly selected, 2 wards per Town council were selected randomly based on probability proportional to size sampling, and 4 villages were selected in each town council. In each village, we selected the first household across the road and selected the rest of the households consecutively. Approximately 40 households were picked from each village in Rubindi-Ruhumba, while about 20 households were selected from each village in Bwizibwera-Rutooma town council. Within each selected household, we followed the eligibility criteria (household heads aged 18 years and older and residents for at least 6 months). Only one participant was selected from each household in case more than one of them was eligible.

The sample size was determined using Krejcie and Morgan’s (1970) method, with a Z-value of 1.96 corresponding to a 95% confidence level, a margin of error of ±5%, and an anticipated response proportion of 80%. Based on a total population of 23,800, the calculated sample size was 240 participants. Proportional allocation was used according to the population distribution of the two town councils: Bwizibwera-Rutooma (32%) contributed 77 participants, while Rubindi-Ruhumba (68%) contributed 163 participants.

### Data collection tool, methods and procedures

The study used a researcher-administered questionnaire developed from similar previous studies to collect data (21). The study tool was pretested among 20 individuals in Ibanda district, and the information obtained was used for adjusting the tool before data collection. The questionnaire was translated into the local language (Runyankore) to cater for participants who lacked proficiency in English. The tool consisted of the sociodemographic characteristics (age, sex, education level, marital status), willingness to pay for solid waste management, and factors associated with willingness to pay for solid waste management. Data was collected using face-to-face interviews with the participants in their households. The interviews were conducted by three trained research assistants who had a Bachelor of Science in Public Health. Research assistants selected the first household across the road and selected the rest of the households consecutively in each village. Each participant was screened for eligibility, consented and recruited into the study. Participants were interviewed in private spaces of their choice. Each interview took approximately 15-20 minutes, and each study tool was checked for completeness after every interview.

### Study variables

The dependent variable in this study was the willingness to pay for solid waste management and was measured using a binary response (yes/no). On the other hand, independent variables included factors like sociodemographic characteristics, such as age in completed years, sex (male/female), highest education level attained, marital status (married/unmarried), and income level (>100,000 Ugx/<100,000 Ugx). Other factors were: quantity of solid waste generated, measured according to the number of polythene bags filled, places of waste disposal, whether waste is dumped in a rubbish pit, collected by service providers or by the town council, and frequency of garbage collection (daily/weekly/monthly).

### Data management and analysis

All digital files were stored encrypted, password-protected and physical records were stored in locked cabinets, only accessible to the research team. A data entry screen was created in Microsoft Excel 2016, and data was entered in duplicates by research assistants. Data cleaning was done, and the data were exported to STATA version 17.0 for analysis.

Continuous Variables were summarized using means and standard deviation, while categorical variables were analyzed using proportions and frequencies. In this study, willingness to pay was analyzed as a dichotomous outcome by calculating the proportion of participants who reported a positive willingness to pay, expressed as a percentage with confidence intervals. The factors associated with “willingness to pay” for solid waste management were analyzed using bivariate and multivariable logistic regression. At bivariate analysis, variables with a p-value <0.25 and had biological plausibility were adopted for multivariable analysis. Model specification was evaluated using the link test to verify that the model was correctly specified. Confounding was assessed using the 10% change in estimate rule by comparing the odds ratios across crude, partially adjusted and fully adjusted models. The Final Model was built using backward conditioning, and factors with a P-value of <0.05 were considered significant at a 95% level of confidence.

### Ethical considerations

The study adhered to the ethical principles according to the Helsinki Declaration. It was reviewed and approved by the Research and Ethics Committee of Mbarara University of Science and Technology (MUST-2024-1781). Administrative clearance was obtained from the Mbarara District Chief Administrative Officer before data collection. Written informed consent was obtained from all the study participants, and they were assured of maximum confidentiality and protection. Participants’ identities were maintained anonymous with unique codes instead of their names.

## Results

### Sociodemographic characteristics of the study participants

Of the 250 participants enrolled in this study, the majority, 61.2% (153/250), were female. The mean age of the study participants was 37.63±11.8 years. More than half, 57.6% (144/250) of the participants were aged 18-38 years, and most, 80.4% (201/250) of the participants had some form of formal education. Regarding the amount of solid waste generated, the majority, 36% (90/250), produced 1-2 polythene bags of solid waste per week,and half, 51.6% (129/250), reported that solid waste was collected weekly from their households (**Table 1**).

**Table 1:** Sociodemographic characteristics of adult residents in households of Mbarara district, southwestern Uganda, March 2025.

| Variable | Categories | Frequency (%) | Willingness to Pay |  |
| --- | --- | --- | --- | --- |
|  |  |  | No n(%) | Yes n (%) |
| Sex | Female | 153 (61.2) | 65(42.5) | 88(57.5) |
|  | Male | 97 (38.8) | 29 (29.9) | 68(70.1) |
| Age (in years)<br>Mean ± SD =<br>37.6±11.8 | 18-38 years | 144 (57.6) | 61(42.4) | 83 (57.6) |
|  | 39-75 years | 106 (42.4) | 33 (31.1) | 73 (68.9) |
| Education level | No formal education | 51 (20.4) | 22 (43.1) | 29(56.9) |
|  | Primary | 102 (40.8) | 39 (38.2) | 63(61.8) |
|  | Secondary | 74 (29.6) | 25 (33.8) | 49(66.2) |
|  | Tertiary | 23 (9.2) | 8 (34.8) | 15(65.2) |
| Marital Status | Unmarried | 49 (19.6) | 21 (42.9) | 28 (57.1) |
|  | Married | 201(80.4) | 73(36.3) | 128 (63.7) |
| House Ownership | Rent | 173 (69.2) | 72 (41.6) | 101 (58.4) |
|  | Self-owned | 77 (30.8) | 22 (28.6) | 55 (71.4) |
| Monthly Income (Ugandan Shillings) | <100,000 | 93 (37.2) | 46 (49.5) | 47(50.5) |
|  | >100,000 | 157 (62.8) | 48 (30.6) | 109 (69.4) |
| Quantity of solid wastes generated | Less than 1 Polythene bag | 60 (24.0) | 31 (51.7) | 29 (48.3) |
|  | 1-2 Polythene bags | 90 (36.0) | 28 (31.1) | 62 (68.9) |
|  | 2-8 Polythene bags | 41 (16.4) | 16 (39.0) | 25 (61.0) |
|  | More than 9 polythene bags | 59 (23.6) | 19 (32.2) | 40 (67.8) |
| Places of waste disposal | Collected by service providers | 9 (3.6) | 7 (77.8) | 2 (22.2) |
|  | Rubbish pit in the back yard | 64 (25.6) | 36 (56.2) | 28 (43.8) |
|  | Garbage dumping site | 82 (32.8) | 21 (25.6) | 61 (74.4) |
|  | Town Council Skip | 95 (38.0) | 30(31.6) | 65 (68.4) |
| Frequency of collection of solid waste by service providers | Daily | 89 (35.6) | 38 (42.7) | 51 (57.3) |
|  | Weekly | 129 (51.6) | 40 (31.0) | 89 (69.0) |
|  | Monthly and not regularly collected | 32 (12.8) | 16 (50.0) | 16 (50.0) |

*SD: Standard Deviation, 100,000ugx≈28 USD*

### Willingness to pay for solid waste management services

Out of the 250 participants enrolled in this study, 62% (156/250; 95% C.I.: 56.2%-68.2%) were willing to pay for solid waste management services. Of the 156 respondents who were willing to pay, the majority, 64.1% (100/156), were willing to pay one thousand Uganda shillings or more, while a small proportion, 4.5% (7/156), were willing to pay less than five hundred Uganda Shillings for solid waste management services.

Among the participants willing to pay for better solid waste management services, the most frequently cited reason that motivated participants to pay was keeping the environment clean and tidy, 59.0% (92/156), followed by health and disease prevention, reported 35.9% (56/156). Other notable reasons included conserving the environment, lack of nearby disposal grounds, and adherence to regulatory requirements.

### Factors associated with willingness to pay for solid waste management

The association of independent variables with willingness to pay for solid waste management services was investigated using both bivariate and multivariable logistic regression. At bivariate analysis, gender (cOR = 1.7, 95% CI: 1.0-3.0; *p*-value = 0.046), monthly income greater than 100,000 Ug Shillings (cOR = 2.2, 95% CI: 1.3-3.8; *p*-value = 0.003), waste generation of 1-2 bags (cOR = 2.4, 95% CI: 1.2-4.7; *p*-value = 0.012), waste generation of more than 9 bags (cOR = 2.3, 95% CI: 1.1-4.9; *p*-value = 0.033), garbage dumping site as the place of waste disposal (cOR = 10.2, 95% CI: 2.0-52.8; *p*-value = 0.006), town council skip as the place of waste disposal (cOR = 7.6, 95% CI: 1.5 -38.7; *p*-value = 0.015), and weekly frequency of waste collection (cOR = 2.22, 95% CI: 1.01-4.89; *p*-value = 0.046) were significant.

In multivariable logistic regression, four factors remained statistically significant. Male participants had 2.4 times higher odds of being willing to pay for solid waste management services (aOR = 2.4, 95% CI: 1.2-4.6; *p*-value = 0.011) compared to females. Participants with a monthly income greater than 100,000 Ug Shillings had 2.2 times higher odds of being willing to pay (aOR = 2.2, 95% CI: 1.7-4.1; *p*-value = 0.015) compared to those with less than 100,000 Ug Shillings. Participants who utilize town council services for solid waste disposal had 7.75 times higher odds of being willing to pay (aOR = 7.75, 95% CI: 1.35-44.47; *p*-value = 0.022) compared to those whose waste is collected by private service providers. Participants receiving weekly waste collection were significantly more willing to pay than those with monthly or irregular collection (aOR = 2.62; 95% CI: 1.06-6.50; *p*=0.038) (**Table 2**).

**Table 2:**
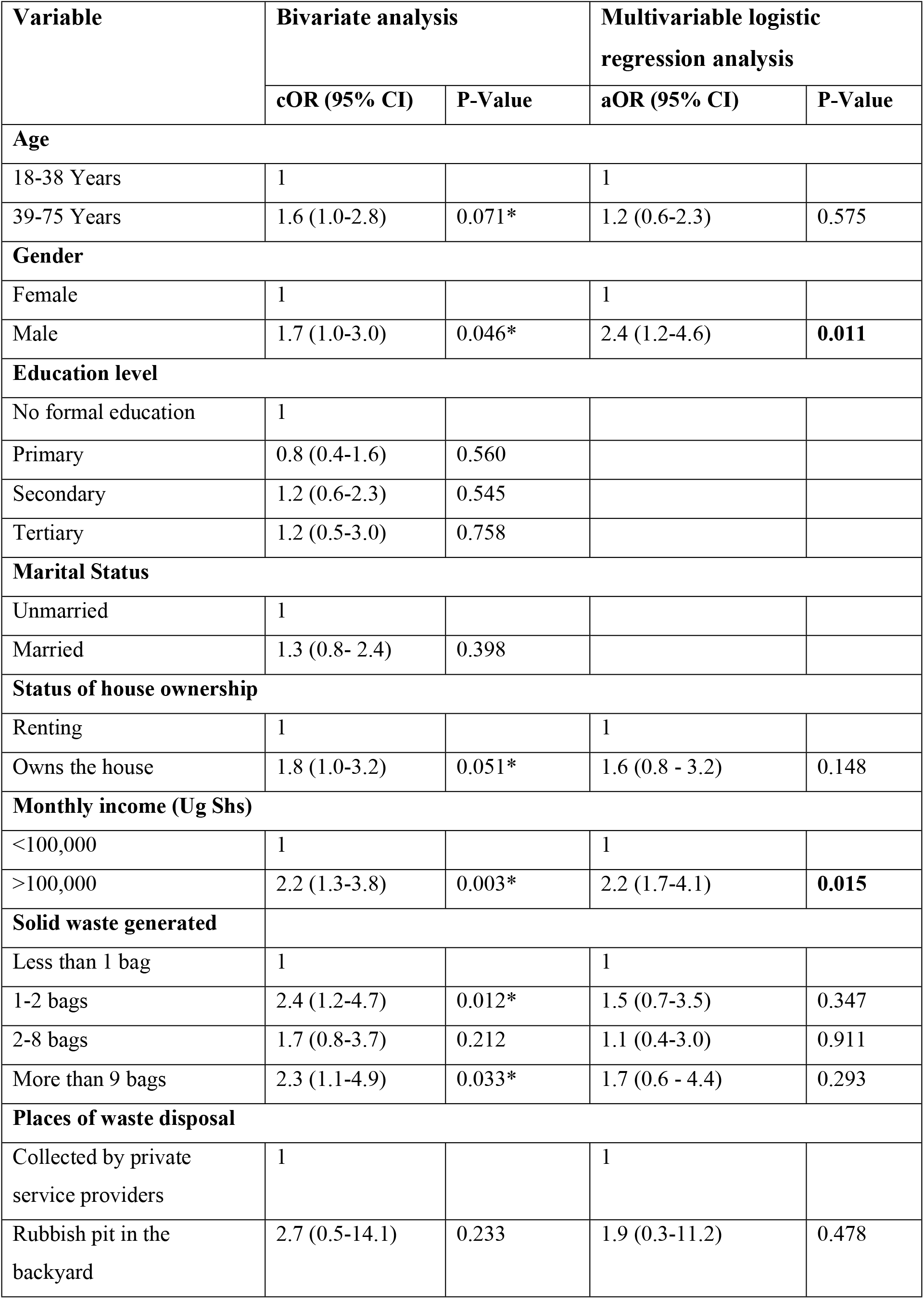
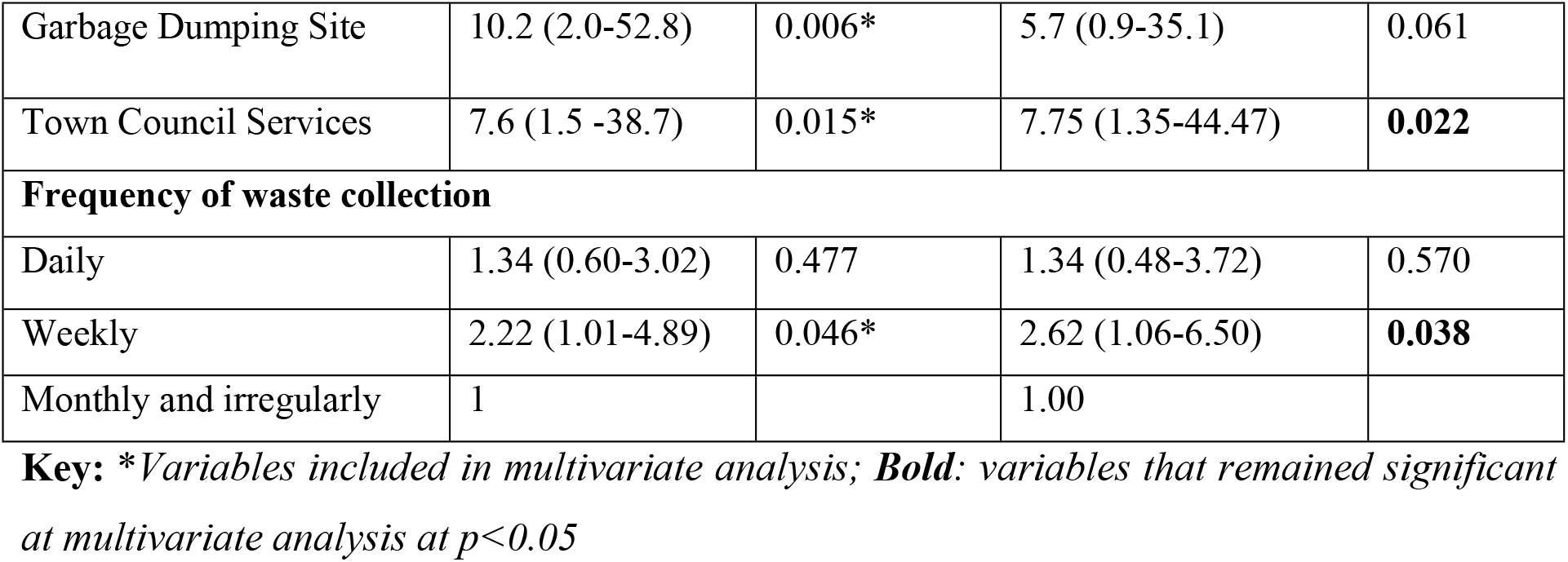
Bivariate and multivariable logistic regression of the factors associated with willingness to pay for waste management services among households in Mbarara district, southwestern Uganda, March 2025.

| Variable | Bivariate analysis |  | Multivariable logistic regression analysis |  |
| --- | --- | --- | --- | --- |
|  | cOR (95% CI) | P-Value | aOR (95% CI) | P-Value |
| **Age** |  |  |  |  |
| 18-38 Years | 1 |  | 1 |  |
| 39-75 Years | 1.6 (1.0-2.8) | 0.071* | 1.2 (0.6-2.3) | 0.575 |
| **Gender** |  |  |  |  |
| Female | 1 |  | 1 |  |
| Male | 1.7 (1.0-3.0) | 0.046* | 2.4 (1.2-4.6) | **0.011** |
| **Education level** |  |  |  |  |
| No formal education | 1 |  |  |  |
| Primary | 0.8 (0.4-1.6) | 0.560 |  |  |
| Secondary | 1.2 (0.6-2.3) | 0.545 |  |  |
| Tertiary | 1.2 (0.5-3.0) | 0.758 |  |  |
| **Marital Status** |  |  |  |  |
| Unmarried | 1 |  |  |  |
| Married | 1.3 (0.8- 2.4) | 0.398 |  |  |
| **Status of house ownership** |  |  |  |  |
| Renting | 1 |  | 1 |  |
| Owns the house | 1.8 (1.0-3.2) | 0.051* | 1.6 (0.8 - 3.2) | 0.148 |
| **Monthly income (Ug Shs)** |  |  |  |  |
| <100,000 | 1 |  | 1 |  |
| >100,000 | 2.2 (1.3-3.8) | 0.003* | 2.2 (1.7-4.1) | **0.015** |
| **Solid waste generated** |  |  |  |  |
| Less than 1 bag | 1 |  | 1 |  |
| 1-2 bags | 2.4 (1.2-4.7) | 0.012* | 1.5 (0.7-3.5) | 0.347 |
| 2-8 bags | 1.7 (0.8-3.7) | 0.212 | 1.1 (0.4-3.0) | 0.911 |
| More than 9 bags | 2.3 (1.1-4.9) | 0.033* | 1.7 (0.6 - 4.4) | 0.293 |
| **Places of waste disposal** |  |  |  |  |
| Collected by private service providers | 1 |  | 1 |  |
| Rubbish pit in the backyard | 2.7 (0.5-14.1) | 0.233 | 1.9 (0.3-11.2) | 0.478 |
| Garbage Dumping Site | 10.2 (2.0-52.8) | 0.006* | 5.7 (0.9-35.1) | 0.061 |
| Town Council Services | 7.6 (1.5 -38.7) | 0.015* | 7.75 (1.35-44.47) | **0.022** |
| **Frequency of waste collection** |  |  |  |  |
| Daily | 1.34 (0.60-3.02) | 0.477 | 1.34 (0.48-3.72) | 0.570 |
| Weekly | 2.22 (1.01-4.89) | 0.046* | 2.62 (1.06-6.50) | **0.038** |
| Monthly and irregularly | 1 |  | 1.00 |  |

**Key:** \*Variables included in multivariate analysis; **Bold:** variables that remained significant at multivariate analysis at $p < 0.05$

## Discussion

This study assessed willingness to pay for solid waste management services and associated factors in two selected town councils in Mbarara district. Close to two-thirds, 62% (156/250), of the study participants were willing to pay for solid waste management, and the factors associated with willingness to pay include male gender, earning more than 100,000 Ugandan shillings, weekly waste collection, and using town council services for waste collection.

Our findings indicate that 62% of participants were willing to pay for solid waste management services, a proportion that aligns closely with similar studies such as 63% in Tanzania (19), 64.4% in Nigeria (17), 61% in Sri Lanka (26), and 64% in Kampala, Uganda (21). This consistency suggests a general recognition across diverse settings of the importance of investing in waste management services. However, our observed willingness to pay is higher than the 48% reported in Lira City (20) and the 53.7% from a multi-city study in Ghana (27), possibly reflecting differences in economic conditions. Semi-urban settings like Mbarara may have relatively lower living costs and less financial pressure, making it easier for households to allocate funds for waste services. In contrast, residents of larger urban centres may experience higher financial burdens, which could limit their ability or willingness to pay for such services (28). On the other hand, our findings fall below the 83% reported in Nepal (29) and the 86.3% in Ethiopia (30), where studies employed the contingent valuation method, an approach known to prompt more favourable responses toward payment. This methodological difference could partly explain the higher willingness observed in those contexts. Nonetheless, the 62% WTP identified in our study demonstrates a substantial level of public support for structured waste management systems in Mbarara District. It presents a valuable opportunity for local governments and private stakeholders to design and implement sustainable, cost-recovery models for SWM services. Additionally, with 64.1% of participants willing to pay UGX 1,000 (approximately USD 0.30) or more for these services, there is potential to introduce affordable service fees. However, any payment model must remain sensitive to the socio-economic realities of the population to ensure equity, accessibility, and long-term sustainability.

We found that those male participants were 2.4 times more likely to pay for solid waste management services compared to female participants. Our finding is consistent with those obtained in studies done in Ethiopia(31) and China (32). On the contrary, previous studies reported female gender as being associated with willingness to pay for solid waste management services (30, 33). Additionally, the proportion of men earning more than 100,000 Ugandan shillings was 69.1% (67/97) higher than that of women at 58.8% (90/153). This means that men hold greater financial control within households in our setting, leading to a higher reported WTP for solid waste management services. Furthermore, men might have a different perception of the socioeconomic benefits associated with improved SWM as previously reported in literature (34). Since men are the majority heads of households, this finding is key in guiding the design and implementation of sustainable Solid waste management strategies in Mbarara District. Additionally, there is need to understand why women were less likely to pay for Solid waste management services so that strategies will be inclusive.

Participants with a monthly income greater than 100,000 Ugandan Shillings (approximately 28 USD) had 2.2 times higher odds of willing to pay compared to those with less than 100,000 Ugandan Shillings. This is consistent with previous findings where increased income is a significant factor (20, 26, 29, 33). In the context of waste management, households with high income may value services that prevent potential health risks and improve community cleanliness (34). This highlights an opportunity for local governments and private providers to mobilize financial contributions more effectively from middle- and higher-income segments, who are both willing and able to pay. As such, it is important to tailor fee structures to reflect these income differences to ensure affordability and maximize revenue generation for sustainable service provision.

In this study, participants who disposed of their waste through town council services had nearly eight times higher odds of being willing to pay for solid waste management services compared to those whose waste was collected by private providers. This finding is consistent with previous research suggesting that public waste services are often more widespread and perceived as more legitimate or community-oriented, fostering a stronger sense of shared responsibility and public trust (14, 35). Town council services may also be more accessible to a wider socioeconomic spectrum, particularly in semi-urban settings where private waste management options are limited or more expensive. In contrast, the use of private providers is frequently observed in areas underserved by public systems or among higher-income households who can afford individualized services (36). This disparity in WTP based on service provider highlights the importance of developing an inclusive and equitable SWM strategy, one that ensures both accessibility and affordability across diverse communities. Strengthening public waste services, alongside regulated collaboration with private actors, may enhance public willingness to financially support sustainable waste management systems.

Study findings show a positive association between the frequency of garbage collection and the participants’ willingness to pay for SWM services. Participants receiving weekly waste collection were nearly 3 times more willing to pay than those whose waste was collected on a monthly or irregular basis. This association suggests that the perceived value and reliability of waste management services significantly influence household decisions to financially support them. Regular and predictable collection not only reduces the health and environmental hazards associated with waste accumulation but also builds public trust and satisfaction with the service, thereby increasing the likelihood of cost-sharing participation (33, 37). The higher WTP for this consistent service shows that residents are likely willing to contribute more financially when they experience tangible and regular benefits from SWM services. As such, local authorities and private waste managers should prioritize regular service delivery as a strategic lever to foster greater willingness among residents to invest in sustainable SWM systems.

This study provides important evidence on the factors associated with willingness to pay for solid waste management in Mbarara District, southwestern Uganda. However, this study has some limitations that need to be accounted for in the interpretation of its findings. The reliance on self-reported data may have introduced response bias, and the cross-sectional design restricts the ability to draw causal inferences. To address these limitations, the study utilized well-trained research assistants and a pre-tested structured questionnaire to ensure consistency and minimize reporting bias.

## Conclusion

Nearly two-thirds of the participants were willing to pay for solid waste management services in this study. Willingness to pay was associated with male gender, monthly income above 100,000 UGX, reliance on town council waste collection services and weekly collection of the solid wastes. The town council leadership needs to prioritize regular investment in solid waste management services to ensure that waste collection and disposal occur at least weekly. Furthermore, the councils should establish an inclusive and flexible payment structure that considers the diverse income levels and payment preferences of town dwellers, as these factors significantly influence willingness to pay.

## Data Availability

All relevant data are within the maniscript and its supporting information files

## Abbreviations

WTP: Willingness to pay
SWM: Solid waste management
UGX: Ugandan shillings
MUST: Mbarara University of Science and Technology
cOR: crude odds ratio
aOR: adjusted odds ratio.

## Acknowledgement

The authors sincerely appreciate the contribution of research assistants and the town council officials of Mbarara district during the implementation of this study.

## Notes

### Competing Interest Statement

The authors have declared no competing interest.

### Funding Statement

The authors received no specific funding for this work

